# Multisystem Inflammatory Syndrome in Children: Survey of Early Hospital Evaluation and Management

**DOI:** 10.1101/2020.07.29.20164459

**Authors:** Matthew L. Dove, Preeti Jaggi, Michael Kelleman, Mayssa Abuali, Jocelyn Y. Ang, Wassim Ballan, Sanmit K. Basu, M. Jay Campbell, Sathish M Chikkabyrappa, Nadine F. Choueiter, Katherine N. Clouser, Daniel Corwin, Amy Edwards, Shira J. Gertz, Rod Ghassemzadeh, Rima J. Jarrah, Sophie E. Katz, Stacie M. Knutson, Joseph D. Kuebler, Jennifer Lighter, Christine Mikesell, Kanokporn Mongkolrattanothai, Ted Morton, Natasha A. Nakra, Rosemary Olivero, Christina M. Osborne, Sarah Parsons, Laurie E. Panesar, Rupal M. Patel, Jennifer Schuette, Deepika Thacker, Adriana H. Tremoulet, Navjyot K. Vidwan, Matthew E. Oster

## Abstract

**Background:** In the absence of evidence-based therapies for Multisystem Inflammatory Syndrome in Children (MIS-C), we aimed to describe the similarities and differences in the evaluation and treatment of MIS-C at hospitals in the United States.

**Methods:** We conducted a cross-sectional survey from June 16 to July 16, 2020 of U.S. children’s hospitals regarding protocols for patients with MIS-C. Elements included hospital characteristics, clinical definition of MIS-C, evaluation, treatment, and follow-up. We summarized key findings and compared results from centers that had treated >5 patients vs. those that had treated ≤5 patients.

**Results:** Forty centers of varying size and experience with MIS-C participated. About half (21/40) of centers required only 1 day of fever for MIS-C to be considered. In the evaluation of patients, there was often a tiered approach. Intravenous immunoglobulin was the most widely used medication to treat MIS-C (98% of centers). Corticosteroids were listed in 93% of protocols for primarily the moderate or severe cases. Aspirin was commonly used including for mild cases, whereas heparin or low molecular weight heparin were used primarily in severe cases. In severe cases, anakinra and vasopressors were frequently recommended. Nearly all centers (39/40) recommended follow up with cardiology. There were similar findings between centers that had treated >5 patients vs. those that had treated ≤5 patients. A supplement containing hospital protocols is provided.

**Conclusion:** There are many similarities yet some key differences between hospital protocols for MIS-C. These findings can help healthcare providers learn from others regarding options for managing MIS-C patients.

*Article Summary:* This survey of U.S. hospitals highlights the interhospital similarities and differences in management of Multisystem Inflammatory Syndrome in Children.

*What’s Known on This Subject:* MIS-C is a novel and life-threatening disease in children associated with COVID-19. Early cases were treated with immunomodulatory agents similar to current guidelines for Kawasaki disease. There are currently no evidence-based guidelines for treatment of MIS-C.

*What This Study Adds:* This study describes the protocolized evaluation and treatment of children with MIS-C at 40 hospitals in the U.S. These findings can help other hospitals create protocols to care for these children at their centers.

## BACKGROUND

In April 2020, physicians in the United Kingdom and France identified an outbreak of children admitted to the pediatric intensive care unit with a hyperinflammatory condition characterized by fever, cardiovascular shock, and suspected SARS-CoV-2 infection: Paediatric Multisystem Inflammatory Syndrome – Temporally Associated with SARS-CoV-2 (PIMS-TS).^1-3^ The Centers for Disease Control and Prevention (CDC) subsequently released a health advisory in May 2020 for Multisystem Inflammatory Syndrome in Children (MIS-C), defining this syndrome as children with fever, laboratory evidence of inflammation, multisystem organ involvement, severe illness, and SARS-CoV-2 infection or exposure.^4^ Other clinical characteristics include acute myocarditis, respiratory failure, features of Kawasaki Disease (KD) and features of toxic shock syndrome.^5^ This rare but life-threatening condition has been reported with increasing frequency in the United States, and growing evidence establishes MIS-C as an immune-mediated condition following SARS-CoV-2 infection.^6-9^

Given the novelty of this new syndrome, evidence-based guidelines for management of children with MIS-C are lacking. Early reports of MIS-C highlight the variability in the evaluation and management of these patients.^2,5-7,9-11^ The American College of Rheumatology and the American Academy of Pediatrics have released guidelines, but these are based primarily on expert opinion.^12,13^ In the absence of evidence-based therapies for MIS-C, many centers have created protocols to guide hospital evaluation and management. The purpose of this study is to describe the similarities and differences in the evaluation and treatment of MIS-C at hospitals in the United States.

## METHODS

We conducted a cross-sectional survey of U.S. children’s hospitals regarding their protocols for patients with MIS-C. Participants were recruited via e-mails to pediatric cardiology and infectious disease list serves and via direct contact to physicians known to be coordinating the MIS-C response at their hospital. The survey was administered from June 16 to July 16, 2020 through the electronic database Research Electronic Data Capture (REDCap) at Children’s Healthcare of Atlanta (CHOA).^14,15^ REDCap is a secure, web-based software platform designed to support data capture for research studies, providing 1) an intuitive interface for validated data capture; 2) audit trails for tracking data manipulation and export procedures; 3) automated export procedures for seamless data downloads to common statistical packages; and 4) procedures for data integration and interoperability with external sources. No patient data were collected as part of this inquiry, and this study was considered Non-Human Subjects Research by the CHOA Institutional Review Board.

We developed an online questionnaire to learn about the protocol at each center (Supplement 1). Elements of the questionnaire included hospital characteristics (location, number of pediatric beds, number of MIS-C patients treated), clinical definition of MIS-C (duration of fever, organ system involvement, evidence of SARS-CoV-2 infection), evaluation (laboratory studies, imaging), treatment (medications and dosages), and follow-up. Finally, participants were invited to share their protocol for inclusion in this publication. Participants at centers without a protocol were able to complete the survey but their responses were excluded from the analyses.

We performed descriptive statistics to summarize quantitative elements via SAS 9.4 and Microsoft Excel. We reviewed the qualitative elements for key themes and summarized the responses as appropriate. We excluded survey responses which did not have sufficient data for analysis. We then performed a subanalysis to compare the quantitative elements comparing the responses of those centers who had treated >5 patients with MIS-C as compared to those centers with ≤5 patients. In the subanalysis we conducted chi-square analyses, or Fisher’s exact test where appropriate. Finally, for the subanalysis we performed a sensitivity analysis comparing results for centers that had treated >10 MIS-C patients vs. those that had treated ≤10 patients.

## RESULTS

There were 48 surveys completed from participants at 48 unique centers across the United States. One record was excluded due to insufficient data submitted, 6 records were excluded because the center did not have a protocol, and 1 record was removed after submission at the request of the contributing center. Thus, survey responses from 40 centers were available for analysis (Figure 1). Protocols from 32 centers were submitted with the survey and are included in Supplement 2.

**Figure 1.**
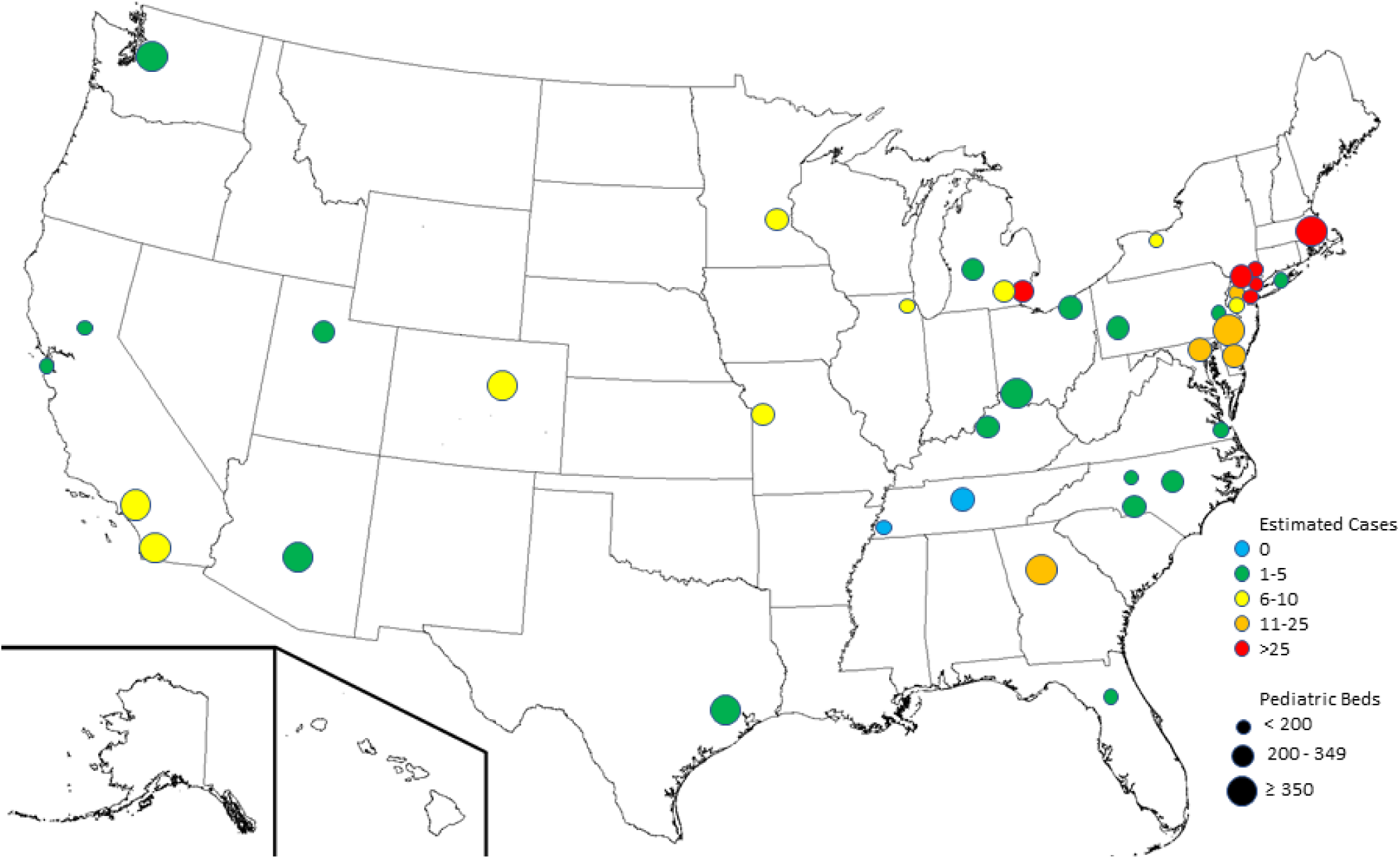
There were 40 hospitals in the United States with protocols for the evaluation and management of Multisystem Inflammatory Syndrome in Children (MIS-C) that responded to the survey. The hospitals varied in both their size (by number of pediatric hospital beds) and their experience in treating patients with MIS-C.

### Hospital characteristics

Participating centers varied in size: 15 small pediatric centers (≤200 pediatric beds), 15 medium centers (200-≤350 pediatric beds), and 10 large centers (≥350 pediatric beds). Experience with treating MIS-C differed between centers: 2 centers with 0 MIS-C patients, 18 centers with 1-5 patients, 9 centers with 6-10 patients, 5 centers with 11-25 patients, and 6 centers with >25 patients. Of the 40 protocols, 21 had been revised since inception.

### Definition of MIS-C

All respondents indicated that fever is required as part of the definition of MIS-C, however the duration and degree of fever varied. About half (21/40) of the centers required only 1 day of fever, 2 centers required at least 2 days, 15 centers required at least 3 days, and 2 centers required at least 4 days of fever. Of the 22 centers that specified a minimum temperature for fever, 20 set 38.0°C as the minimum. Almost all (38/40) centers specified the presence of certain organ system involvement; of these, 3 required only 1 organ system, 31 required at least 2 organ systems, and 4 required at least 3 organ systems involved. In 36 of the 40 protocols, abnormal laboratory markers of inflammation were required to meet MIS-C inclusion criteria. Most centers (31/40) did not require laboratory evidence of current or prior SARS-CoV-2 infection. Instead, prior exposure to someone with COVID-19 in the 4 weeks preceding the onset of symptoms sufficed to meet inclusion criteria. Three centers commented that, given the high prevalence of COVID-19 in their community, the requirement of a known exposure is waived as all children are assumed to have had prior exposure to someone with COVID-19 in the preceding 4 weeks. One center commented that the working definition for MIS-C was too broad, resulting in often unnecessary testing and, in at least one case, delayed diagnosis of perforated appendicitis.

### Evaluation of MIS-C

In the evaluation of patients with possible MIS-C, there was often a tiered approach, with some centers performing initial laboratory tests on all patients and then further tests only on those patients with high suspicion of MIS-C or with relevant symptoms (Figure 2). For the identification of SARS-CoV-2, all centers performed PCR testing from a nasopharyngeal or oropharyngeal sample. Most centers also tested for SARS-CoV-2 antibody in all of their possible MIS-C patients. Routine bloodwork included complete blood count, basic metabolic panel, liver function tests, C-reactive protein, and erythrocyte sedimentation rate. Further bloodwork including investigation for inflammation, cardiac involvement, and abnormal anticoagulation were often performed. Further testing including electrocardiogram, echocardiogram, urinalysis, and chest radiograph were common. Evidence of potential alternative causes or co-infection was routinely pursued via blood culture or respiratory viral panel. For admitted patients, the infectious disease service was almost universally consulted, followed by cardiology, rheumatology, and hematology.

**Figure 2.**
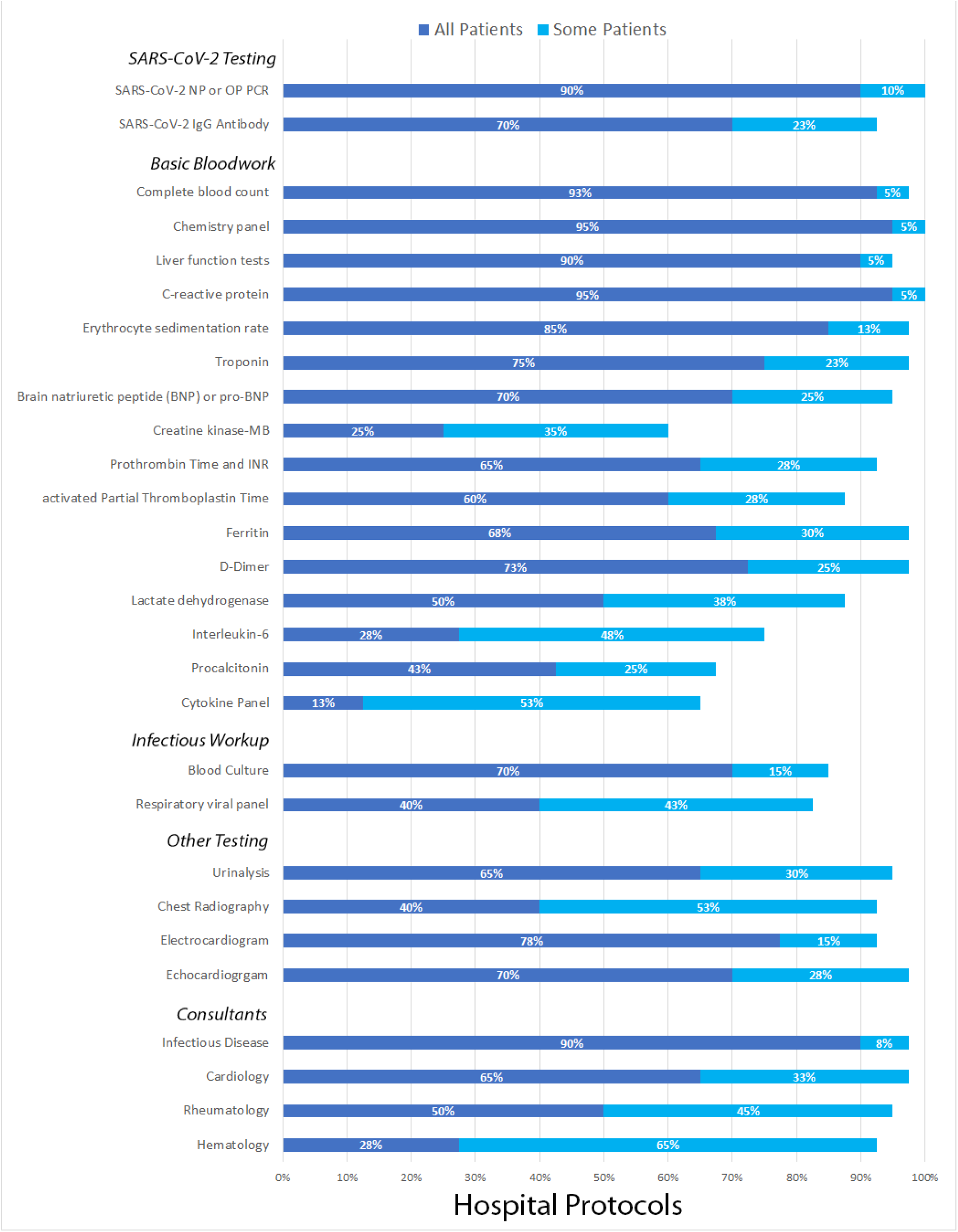
The evaluation of Multisystem Inflammatory Syndrome in Children (MIS-C) varied between centers for SARS-CoV-2 testing, basic bloodwork, infectious workup, ancillary testing, and consultant services. Some protocols included certain aspects for all patients with potential MIS-C, whereas others performed portions for only some patients.

### Treatment of MIS-C

Some centers had a similar treatment approach for all patients, while others varied the approach by severity of illness (Figure 3). Severity of illness was defined specifically at each center, with no uniform definition. Submitted criteria for severity of illness included vasoactive-inotropic score, location in the hospital (intensive care unit vs. general floor), degree of hyperinflammation, and presence of shock or cardiac involvement.

**Figure 3.**
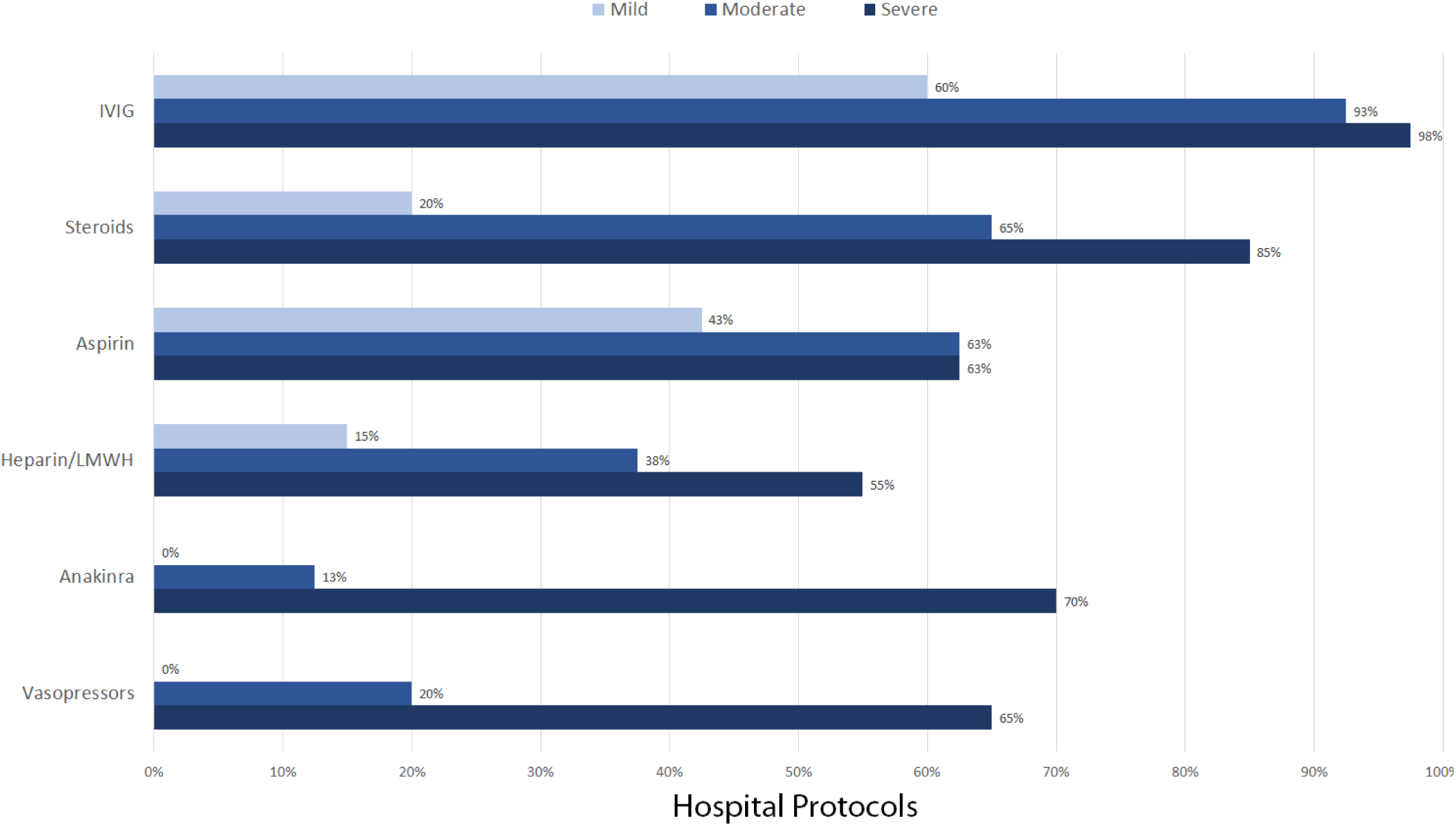
Medical management of Multisystem Inflammatory Syndrome in Children often varied by severity, with severity being defined differently by each center. For centers that gave IVIG, 54% recommended a second dose for patients who were refractory to the first dose. Medications used by ≤20 of the 40 centers are not shown.

Intravenous immunoglobulin (IVIG) was the most widely used medication to treat MIS-C, with 98% of centers including IVIG in their recommendations and 60% recommending the use of IVIG regardless of severity. Of the 39 protocols that mentioned any use of IVIG, 21 recommended a second dose of IVIG for cases that were refractory to the first dose. Corticosteroids were listed in 93% of protocols, although these tended to be reserved primarily for moderate or severe cases. Aspirin was commonly used including the mild cases, whereas heparin or low molecular weight heparin were used primarily in severe cases. In severe cases, anakinra and vasopressors were frequently recommended. Other medications that were recommended in fewer than 25 of the 40 protocols included clopidogrel (3 centers), warfarin (3 centers), remdesivir (10 centers), and tocilizumab or infliximab (13 centers); these medications were primarily reserved for severe or refractory cases. Hydroxychloroquine was not recommended in any of the protocols included in the study.

### Follow-up of patients with MIS-C

While there was no standardized follow-up plan for patients with MIS-C, 26 participants responded that their protocol recommends follow-up similar to that of the American Heart Associations for KD. Nearly all centers (39/40) recommended follow up with cardiology, but they differed as to the timing of follow-up and echocardiogram. Seven centers arranged follow-up in 1 week, 22 centers planned for 2 weeks, and 9 centers in 1 month (with one participant not providing a time of follow-up). Almost all centers (36/40) included aspirin as a discharge medication, with 26 centers including this medicine regardless of degree of coronary involvement. There was no consensus regarding the utility of cardiac magnetic resonance imaging (MRI), with 22 centers mentioning the use of cardiac MRI in MIS-C patients, primarily for evaluation and follow-up of cardiac dysfunction. Protocols at 2 centers included MRI during the initial inpatient hospitalization, 9 centers recommended it during outpatient follow-up (6 at 1-3 months, 3 at 3-6 months), and 10 centers deferred to cardiology regarding when to obtain MRI. Other common specialty follow-up visits included rheumatology by 24 centers, infectious disease by 20, and hematology by 8.

### Subanalysis

In the subanalysis there were similar findings among almost all components of the evaluation and management of MIS-C for centers that had treated >5 patients as compared to those that had treated ≤5 patients (Table 1). The only significant difference was that centers that had treated >5 patients were more likely to arrange follow-up in infectious disease clinic (70% vs. 30%). In the sensitivity analysis comparing findings for those centers that had treated >10 MIS-C patients vs. those that had treated ≤10 patients, there were likewise similar results (data not shown); the only significant difference between centers was that centers that had treated >10 patients were less likely to include anakinra in their protocols (36% vs. 83%, p=0.008).

**Table 1.**
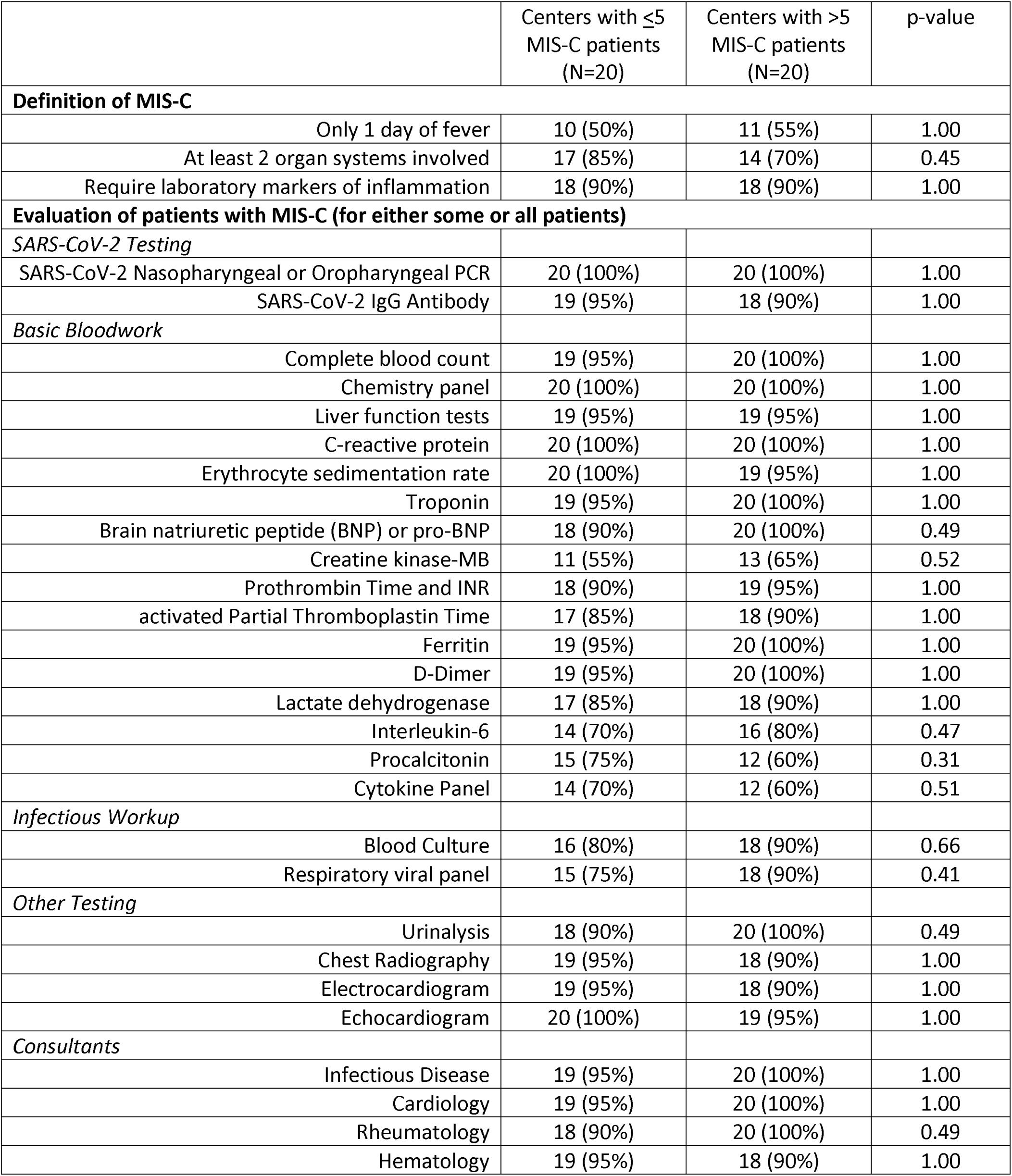
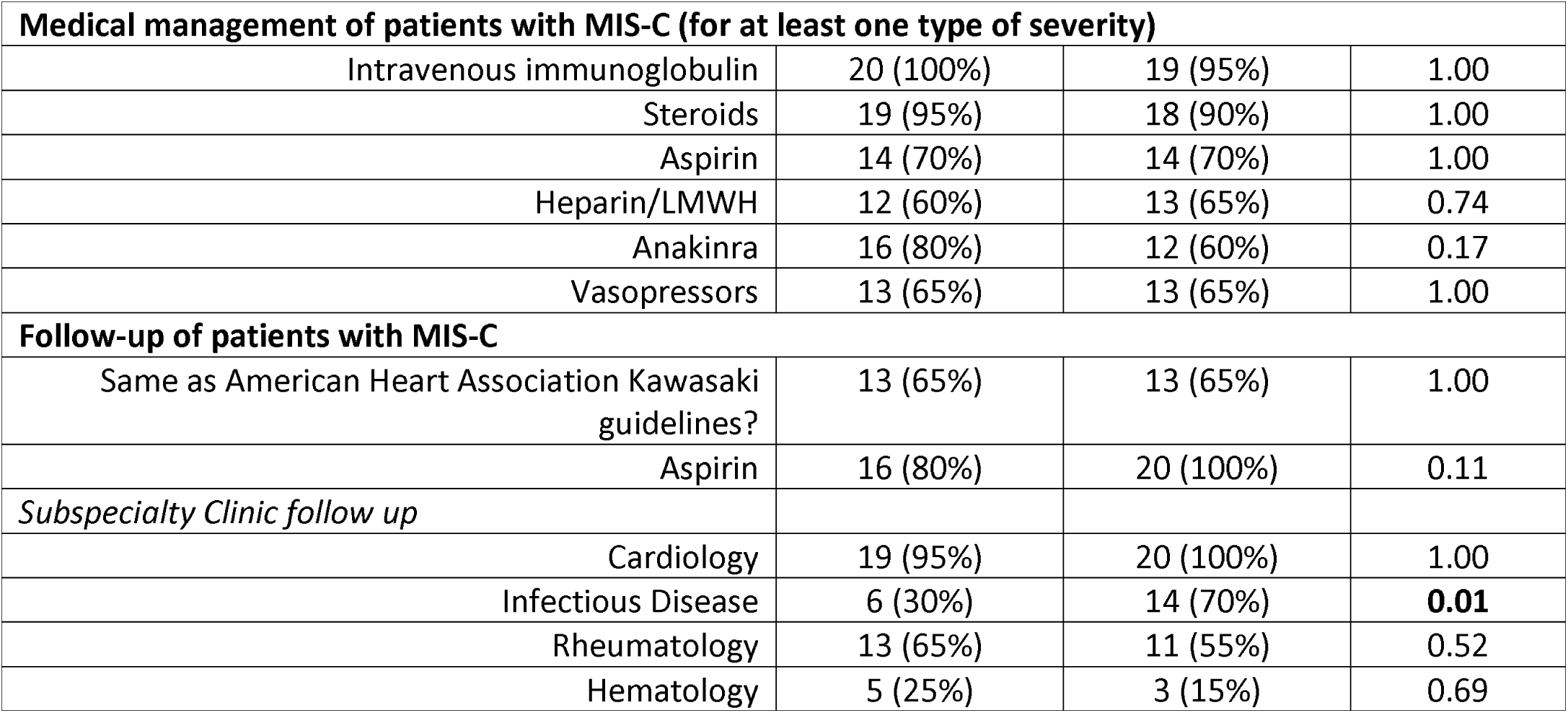
Comparison of evaluation and management of patients with MIS-C based on center experience.

| | Centers with $\leq 5$<br>MIS-C patients<br>(N=20) | Centers with $> 5$<br>MIS-C patients<br>(N=20) | p-value |
| --- | --- | --- | --- |
| **Definition of MIS-C** |  |  |  |
| Only 1 day of fever | 10 (50%) | 11 (55%) | 1.00 |
| At least 2 organ systems involved | 17 (85%) | 14 (70%) | 0.45 |
| Require laboratory markers of inflammation | 18 (90%) | 18 (90%) | 1.00 |
| **Evaluation of patients with MIS-C (for either some or all patients)** |  |  |  |
| *SARS-CoV-2 Testing* |  |  |  |
| SARS-CoV-2 Nasopharyngeal or Oropharyngeal PCR | 20 (100%) | 20 (100%) | 1.00 |
| SARS-CoV-2 IgG Antibody | 19 (95%) | 18 (90%) | 1.00 |
| *Basic Bloodwork* |  |  |  |
| Complete blood count | 19 (95%) | 20 (100%) | 1.00 |
| Chemistry panel | 20 (100%) | 20 (100%) | 1.00 |
| Liver function tests | 19 (95%) | 19 (95%) | 1.00 |
| C-reactive protein | 20 (100%) | 20 (100%) | 1.00 |
| Erythrocyte sedimentation rate | 20 (100%) | 19 (95%) | 1.00 |
| Troponin | 19 (95%) | 20 (100%) | 1.00 |
| Brain natriuretic peptide (BNP) or pro-BNP | 18 (90%) | 20 (100%) | 0.49 |
| Creatine kinase-MB | 11 (55%) | 13 (65%) | 0.52 |
| Prothrombin Time and INR | 18 (90%) | 19 (95%) | 1.00 |
| activated Partial Thromboplastin Time | 17 (85%) | 18 (90%) | 1.00 |
| Ferritin | 19 (95%) | 20 (100%) | 1.00 |
| D-Dimer | 19 (95%) | 20 (100%) | 1.00 |
| Lactate dehydrogenase | 17 (85%) | 18 (90%) | 1.00 |
| Interleukin-6 | 14 (70%) | 16 (80%) | 0.47 |
| Procalcitonin | 15 (75%) | 12 (60%) | 0.31 |
| Cytokine Panel | 14 (70%) | 12 (60%) | 0.51 |
| *Infectious Workup* |  |  |  |
| Blood Culture | 16 (80%) | 18 (90%) | 0.66 |
| Respiratory viral panel | 15 (75%) | 18 (90%) | 0.41 |
| *Other Testing* |  |  |  |
| Urinalysis | 18 (90%) | 20 (100%) | 0.49 |
| Chest Radiography | 19 (95%) | 18 (90%) | 1.00 |
| Electrocardiogram | 19 (95%) | 18 (90%) | 1.00 |
| Echocardiogram | 20 (100%) | 19 (95%) | 1.00 |
| *Consultants* |  |  |  |
| Infectious Disease | 19 (95%) | 20 (100%) | 1.00 |
| Cardiology | 19 (95%) | 20 (100%) | 1.00 |
| Rheumatology | 18 (90%) | 20 (100%) | 0.49 |
| Hematology | 19 (95%) | 18 (90%) | 1.00 |

| **Medical management of patients with MIS-C (for at least one type of severity)** |  |  |  |
| --- | --- | --- | --- |
| Intravenous immunoglobulin | 20 (100%) | 19 (95%) | 1.00 |
| Steroids | 19 (95%) | 18 (90%) | 1.00 |
| Aspirin | 14 (70%) | 14 (70%) | 1.00 |
| Heparin/LMWH | 12 (60%) | 13 (65%) | 0.74 |
| Anakinra | 16 (80%) | 12 (60%) | 0.17 |
| Vasopressors | 13 (65%) | 13 (65%) | 1.00 |
| **Follow-up of patients with MIS-C** |  |  |  |
| Same as American Heart Association Kawasaki guidelines? | 13 (65%) | 13 (65%) | 1.00 |
| Aspirin | 16 (80%) | 20 (100%) | 0.11 |
| *Subspecialty Clinic follow up* |  |  |  |
| Cardiology | 19 (95%) | 20 (100%) | 1.00 |
| Infectious Disease | 6 (30%) | 14 (70%) | **0.01** |
| Rheumatology | 13 (65%) | 11 (55%) | 0.52 |
| Hematology | 5 (25%) | 3 (15%) | 0.69 |

## DISCUSSION

This survey of the protocolized evaluation and treatment of MIS-C across the United States highlights the major similarities and differences among centers; these findings can inform centers creating or modifying MIS-C protocols. Most centers adhered to the MIS-C definition that was put forth by CDC in May 2020. However, some centers require 3 days of fever instead of 1, and centers in areas with high prevalence of COVID-19 do not require positive SARS-CoV-2 test results or a known exposure to someone with the disease. In the evaluation of patients for MIS-C, most centers begin with a tiered approach that is standard for the workup of a febrile illness, with further testing often dictated by symptoms or initial laboratory results. The findings form this survey underscore the collaborative effort to combat MIS-C, as most centers consult multiple subspecialists in the management of these patients. IVIG is a mainstay of treatment at most centers, with corticosteroids, aspirin, and heparin often used as well. Anakinra and vasopressors are frequently used in children with severe illness. Almost all children are discharged on aspirin with planned follow up in cardiology.

Many of the elements of the protocols for MIS-C are similar to those for KD.^2,5-7^ As cases of MIS-C were emerging, the patients were noted to have clinical signs and symptoms of incomplete KD, left ventricular systolic dysfunction as seen in KD shock syndrome, and occasional coronary dilation. The current AHA KD guidelines recommend 2 g/kg IVIG after diagnosis and consideration of a 2-3 week course of tapering corticosteroids for high risk patients. Administration of a second dose of IVIG, high dose IV methylprednisolone, and other immunomodulatory agents are considered if the patient continues to be febrile 36 hours after initial IVIG dose. Low dose aspirin is recommended until 4-6 weeks post onset of illness, and systemic anticoagulation with LMWH or warfarin is recommended for rapidly progressing coronary aneurysms or those with z-score >10.^16^ Our survey revealed that treatment for MIS-C among U.S. children’s hospitals roughly correlated with these recommendations. A large diversion from the KD guidelines was the inclusion of systemic anti-coagulation in some MIS-C protocols. This choice was potentially made due to the elevated d-dimers, frequent deep venous thromboses and pulmonary emboli seen in acutely ill adults with COVID-19, and a small number of reported MIS-C cases with thrombosis.^6,17^ The current choice of therapeutic agents appear reasonable as many patients have recovery of left ventricular systolic function at the time of discharge.^3,5^ Until long-term data are obtained, it is likely reasonable to continue low dose aspirin in the acute 4-6 week period as in KD. However, this approach is not without risk given the concern for Reye syndrome, and the benefit in MIS-C may be less than that in KD as patients with MIS-C are less likely to have elevated platelet counts or coronary involvement ^6,7,10,18^.

The evaluation and management of MIS-C is clearly an evolving process, as more than half of the centers modified their protocols since inception. This iterative process has similarly been seen in the management of adults with COVID-19. For instance, recent data from the RECOVERY trial indicates that dexamethasone may improve mortality in hospitalized adults with severe COVID-19 with severe illness.^19^ It remains to be seen whether such treatment would be useful in children. We anticipate frequent revisions to hospital protocols as new information is learned regarding SARS-CoV-2 and MIS-C.

These findings have important implications during the current pandemic. In the United States, cases of COVID-19 continue to rise, especially among the younger age group.^20^ As a result, we anticipate that cases of MIS-C will rise as well. The findings of this survey can help hospitals with little or no experience yet with these patients to prepare for how to evaluate and manage them. Around the world, the COVID-19 pandemic continues in many countries. In some developing nations, certain treatment options such as IVIG are not readily available. These survey results can help identify other potential options in resource-limited settings.

This study is not without its limitations. First, we did not provide a definition for severity of illness, as severity can differ between institutions. Therefore, what may be considered a moderate case at one center may be severe at another; this difference should be acknowledged when interpreting the treatment options. Second, there was a wide variation in experience in managing MIS-C; there were 6 centers with experience treating >25 patients, and 2 centers with no experience treating MIS-C patients. Thus, some protocols may be based on experience whereas others may be based on personal opinion. We attempted to overcome this limitation by comparing the protocols at those centers with more experience vs. those without. Finally, it is important to recognize that this study captures what centers have recommended for the evaluation and management of MIS-C at their institution, not what has actually been done for those patients. Indeed, protocols may serve as a framework for managing patients with MIS-C, but care may be individualized as dictated by patient signs, symptoms, and response to treatment.

## CONCLUSIONS

MIS-C is a new syndrome with rapidly evolving strategies for evaluating and managing the condition. There are many similarities yet key differences between hospital protocols. In the absence of evidence-based guidelines, these findings can help healthcare providers learn from others regarding options for evaluating and managing patients with MIS-C. It is expected that the understanding of this condition will continue to evolve as more is learned regarding MIS-C and as evidence mounts as to what treatment strategies may be best.

## Data Availability

Data available on request

## Acknowledgements

We would like to thank the additional individuals who contributed data to this survey, including: Eva Cheung MD (NewYork-Presbyterian Morgan Stanley Children’s Hospital of Columbia Irving Medical Center), Lauren Henderson MD MMSc (Boston Children’s Hospital), Whitnee J. Hogan MD (UCSF Benioff Children’s Hospital), Sean Lang MD (Cincinnati Children’s Hospital Medical Center), Jennifer Schuster MD MSCI (Children’s Mercy Kansas City), Renata Shih MD (Congenital Heart Center, University of Florida), Dongngan T. Truong MD MS (University of Utah/Primary Children’s Hospital), Rajiv Verma MD (Children’s Hospital of New Jersey), and Justin P. Zachariah MD MPH (Texas Children’s Hospital, Baylor College of Medicine). We would also like to thank the multidisciplinary teams that helped to create MIS-C protocols at the children’s hospitals in this study, without whose collaborative efforts this study would not have been possible.

## Abbreviations

AHA: American Heart Association
CDC: Centers for Disease Control and Prevention
CHOA: Children’s Healthcare of Atlanta
COVID-19: Coronavirus Disease 2019
IVIG: intravenous immunoglobulin
KD: Kawasaki Disease
MIS-C: Multisystem Inflammatory Syndrome in Children
MRI: magnetic resonance imaging
PCR: polymerase chain reaction
PIMS-TS: Paediatric Multisystem Inflammatory Syndrome - Temporally Associated with
SARS-CoV-2: severe acute respiratory syndrome coronavirus 2
REDCap: Research Electronic Data Capture

## Author Contributions

Dr. Dove and Dr. Oster conceptualized and designed the study, analyzed and interpreted the data, drafted the initial manuscript, critically reviewed and revised the manuscript, and approved of the final manuscript as submitted.

Mr. Kelleman carried out statistical data analysis, critically reviewed and revised the manuscript, and approved of the final manuscript as submitted.

Dr. Jaggi contributed to the study conception, critically reviewed interpretation of the data, reviewed and revised the manuscript, and approved of the final manuscript as submitted.

Dr. Abuali, Dr. Ang, Dr. Ballan, Dr. Basu, Dr. Campbell, Dr. Chikkabyrappa, Dr. Choueiter, Dr. Clouser, Dr. Corwin, Dr. Edwards, Dr. Gertz, Dr. Ghassemzadeh, Dr. Jarrah, Dr. Katz, Dr. Knutson, Dr. Kuebler, Dr. Lighter, Dr. Mikesell, Dr. Mongkolrattanothai, Dr. Morton, Dr. Nakra, Dr. Olivero, Dr. Osborne, Dr. Parsons, Dr. Panesar, Dr. Patel, Dr. Schuette, Dr. Thacker, Dr. Tremoulet, and Dr. Vidwan coordinated acquisition of data, critically reviewed and revised the manuscript, and approved of the final manuscript as submitted.

All authors approved the final manuscript as submitted and agree to be accountable for all aspects of the work.

Supplement 1: Survey for the Treatment of Multisystem Inflammatory Syndrome in Children

Supplement 2: Submitted hospital-based protocols for the treatment of Multisystem Inflammatory Syndrome in Children

